# Designing infection prevalence and seroprevalence surveys for learning infection dynamics

**DOI:** 10.64898/2026.07.01.26356511

**Authors:** Richard Creswell, Nick Golding, Gerard E. Ryan, Oliver Eales, David J. Price, James M. McCaw, Freya M. Shearer

**Affiliations:** Centre for Epidemiology & Biostatistics, Melbourne School of Population and Global Health, The University of Melbourne, Parkville, Victoria, Australia; The Kids Research Institute Australia, Nedlands, Western Australia, Australia; School of Physics, Mathematics and Computing, University of Western Australia, Nedlands, Western Australia, Australia; UWA Centre for Child Health Research, University of Western Australia, Nedlands, Western Australia, Australia; School of Mathematics and Statistics, The University of Melbourne, Parkville, Victoria, Australia; Department of Infectious Diseases, University of Melbourne, at the Peter Doherty Institute for Infection and Immunity, Parkville, Victoria, Australia

**Author notes:** 207 Bouverie St, Parkville VIC 3053 Australia.

**Keywords:** disease surveillance, seroprevalence, infection prevalence, experimental design, time series

## Abstract

Knowledge of the true number of infections over time is valuable for accurately predicting the future course of an epidemic and planning effective interventions, but the number of cases reported offers only a noisy underestimate of the true number of infections. Disease surveillance strategies based on assessing subsets of the population for current infection (infection prevalence surveys) or antibody presence (seroprevalence surveys) yield crucial information about the number truly infected, but are expensive. To explore impact of survey design considerations—both sample size and sampling frequency—on inference of the number of incident infections over time, we coupled agent-based simulations of respiratory virus epidemics with simulations of infection prevalence and seroprevalence surveys. While returns diminish with increased sample size, we find inference generally improved by increasing survey frequency relative to participants-per-round for any given sample size. After survey rounds reach a sufficient frequency, comparable inference performance may be achieved with either more frequent rounds or more participants per round. Rolling designs with tests conducted each day tend to outperform designs in which testing is divided into discrete rounds. We also show that misspecified assumptions about seroreversion may substantially decrease the quality of inference results.

## 1 Introduction

When an infectious disease spreads through a population, the true number of infections arising over time is not directly observable. Reported cases, if available, almost always underestimate the true number of infections ^14^; numerous factors contribute to this underestimation, including asymptomatic infections ^18;3^, people choosing not to test ^16;27^, and limited diagnostic and testing resources ^4^.

However, the true number of infections contains valuable information about an epidemic. Knowledge of the number of infections is crucial to inferring the proportion of the population that is likely to be susceptible to infection, which is valuable for making predictions of future cases, hospitalizations and deaths, or estimating the impact of vaccinations and other interventions ^42;40^. Infection data also provide a less biased, more stable denominator for assessments of symptoms, clinical severity, risk factors, and intervention effectiveness compared to case data ^15;33;34;7;13;49;46;41^.

Infection prevalence surveys, in which a sample of people from the population is tested for current infection, as well as seroprevalence surveys, in which members of a sample are tested for the presence of antibodies indicating a history of infection (or vaccination), are key surveillance strategies for obtaining information about the actual number of infections ^24;45;20;31^. During the COVID-19 pandemic, such surveys were deployed in various regions of the world including, for example, Panama ^21^; Gold Coast, Australia ^48^; Tanzania ^26^; Indiana, Connecticut, Massachusetts, and New York City in the United States of America ^25;11;50;23;12;43^; and the United Kingdom (UK) ^32;36^.

Surveillance based on repeated random surveys took place on a particularly intensive scale in the UK, where infection prevalence and seroprevalence surveys were conducted as part of two major studies: the Real Time Assessment of Community Transmission (REACT), based on repeated cross-sectional surveys of individuals registered with the National Health Service in England; and the Coronavirus Infection Survey (CIS), based on longitudinal sampling of private residential households in the UK. The outputs of these national-scale infection prevalence and seroprevalence surveys in the UK played a central role in understanding transmission, disease severity, risk factors, and intervention effectiveness, and informing policy ^9;47;37^. Despite the utility of infection prevalence and seroprevalence surveys, these modes of disease surveillance can be expensive to conduct (e.g., ^30^). Such expenses are likely to be considered unaf-fordable for many countries, or future events, particularly for the surveillance of endemic pathogens. Thus, designing surveys whose outputs are most useful for public health tasks, at minimal cost, is an important goal. For tasks such as learning the true number of infections arising over time, the outputs of infection prevalence and seroprevalence surveys are used as inputs to a nontrivial inference procedure. Determining the uncertainty in the output of such an inference procedure as a function of survey design parameters, such as sample sizes and frequency of survey rounds, is analytically intractable and therefore requires simulation-based analyses. Hence conventional power analyses for designing infection prevalence and seroprevalence surveys are inadequate and epidemic model-based approaches, e.g., ^5;22^, offer a suitable alternative.

In the present paper, we assume that random surveys consist of infection surveys, seroprevalence surveys, or a combination of both. We focus on key characteristics of the design of an infection prevalence or seroprevalence survey, such as the number of people to include in each random sample, and the times at which to collect the samples. Using simulated epidemic and surveillance data, we investigate which survey designs may be more or less efficient at informing estimates of the true number of infections over time.

## 2 Methods

We first describe the methods by which we generate simulated epidemics and surveys. Next, we describe the methods by which we assess the effectiveness of simulated survey outputs at informing estimates of the true number of infections arising over time. Our study involves repeating this procedure for a multitude of candidate survey designs, thus gaining insights into which designs are more or less effective. A schematic overview of our approach is given in Fig. 1.

**Figure 1:**
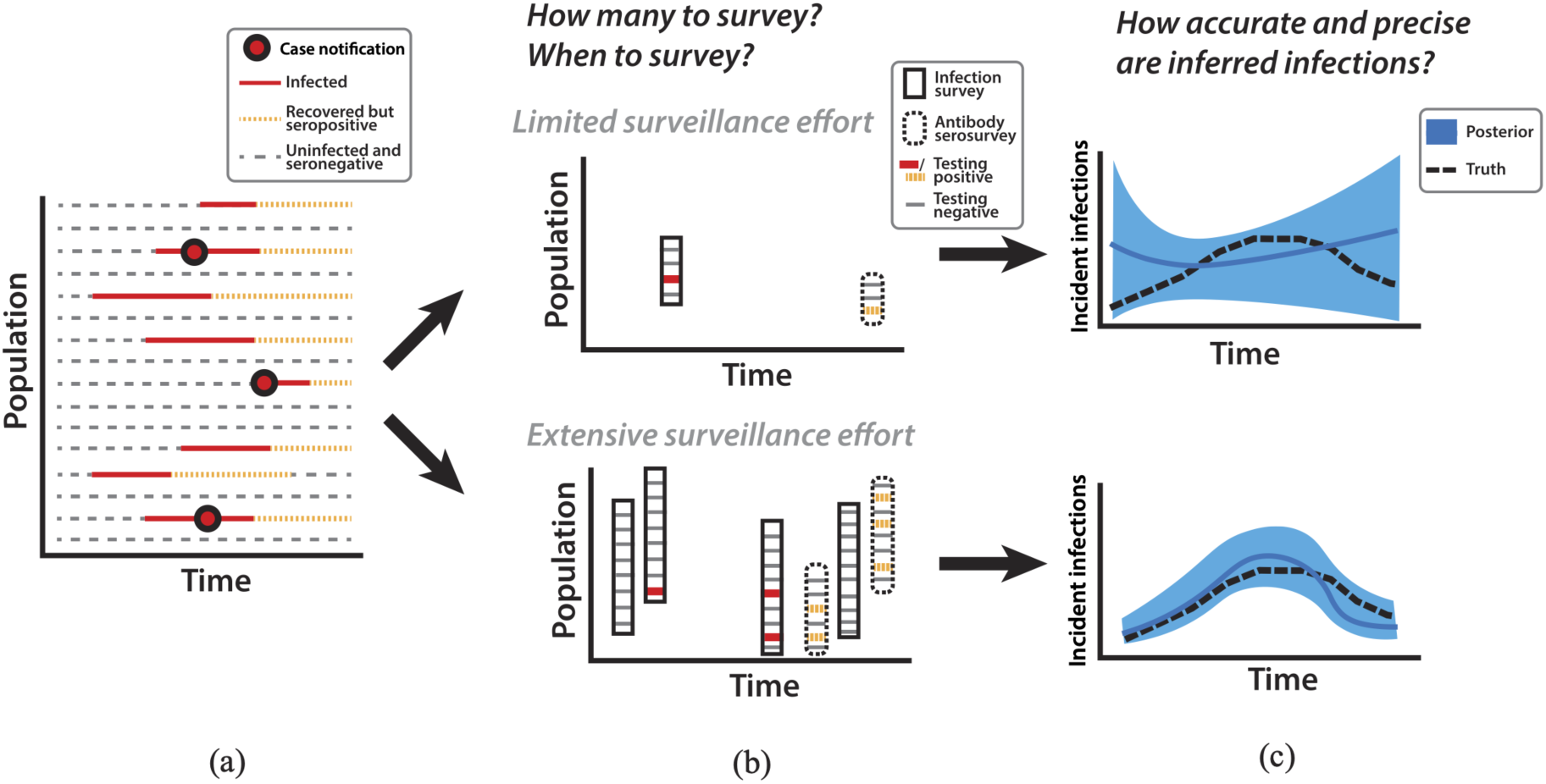
Schematic overview of disease surveillance design for learning infection dynamics. (a) We visualize individuals in the population as horizontal lines; some individuals experience infection and subsequent temporary seropositivity. Some infected individuals may be reported as cases, but many are not. (b) Surveillance activities, consisting of infection prevalence surveys or antibody seropositivity surveys undertaken at different frequencies and with different amounts of sampling, reveal information about the proportion of the population testing positive at the times when surveys are conducted. (c) Limited surveillance efforts fail to precisely inform the true number of infections arising over time, but extensive surveillance effort can lead to more precise estimates.

### 2.1 Simulation of epidemic and surveillance data

#### 2.1.1 Epidemic model

We generated synthetic epidemics using a discrete-time stochastic agent-based model based on the SIRS (Susceptible-Infectious-Recovered-Susceptible) paradigm. This model has *N* agents, each representing an individual person in the population. At each time, each agent belongs to one of three possible statuses: susceptible, infectious, and recovered. At each time, susceptible agents may become infectious according to a probability which depends upon the number of currently infectious agents and a time-varying transmission rate *ρ*(*t*), which is a global parameter of the model. The model simulates progression of infectious individuals to recovered (where recovered individuals are assumed immune and no longer infectious), and progression of recovered individuals back to susceptible, by randomly sampling the number of days each individual remains infectious or recovered according to discretized probability distributions (gamma distribution), whose mean and standard deviation parameters are provided as inputs to the model. Full details of the epidemic simulation model are provided in §S2.

#### 2.1.2 Observation processes

Given an underlying epidemic as described in §2.1.1, we simulated two forms of surveillance data: infection prevalence surveys and seroprevalence surveys.

We assumed that each survey within a series of infection prevalence or seroprevalence surveys is characterized by three parameters: a number of individuals to include in the sample *M*, a start time *t*^start^, and a survey duration (in days) *w*. To simulate one survey, we randomly selected *M* individuals from the population, without replacement. *M/w* of the randomly selected individuals are tested on each day within [*t*^start^*, t*^start^ + *w -* 1]. To avoid the necessity of modelling the duration of test positivity separately from the duration of infectiousness, we assumed that the test for infection (within an infection prevalence survey) targets those (and only those) who are infectious. Likewise, to simulate a seroprevalence survey, we assumed that the test for antibody positivity targets those who are either infectious or recovered. For all results presented in the main text, if the individual had the target status at their sampling time, we simulated a positive test result; if the individual did not have the target status, we simulated a negative test result (i.e., we assumed that the test has perfect sensitivity/specificity for the target status). In §S4 we present additional simulation-based results as well as mathematical analysis for the situation where tests have imperfect sensitivity and specificity.

### 2.2 Configuration of a yearlong epidemic with two waves

The parameter settings of our model (used in results §3.2–§3.3) were selected in order to simulate a yearlong epidemic with two waves. We selected parameter values typical of a real-world respiratory pathogen (e.g., SARS-CoV-2) spreading in a small country, with no prior immunity (i.e., the early phase of a pandemic), although we did not attempt to precisely parametrize any particular epidemic scenario.

We set a total population size *N* = 1 million, and an initial number of infectious individuals, *I*(*t* = 0) = 50, chosen to avoid strong stochastic effects at the beginning of the epidemic. We set transmission rate *ρ*(*t*) according to a time-varying function shown in Figure 2, including a period of lower transmission between days 100 and 150, mimicking the imposition of stringent social restrictions, followed by a resurgence in the transmission rate (which helps induce a second wave of infections) at day 200, reflecting relaxation of social restrictions. We modelled the duration individuals remain infectious using a gamma distribution with mean 7 days and standard deviation 2 days. We modelled the duration individuals remain with recovered status using a gamma distribution with mean 300 days and standard deviation 100 days.

**Figure 2:**
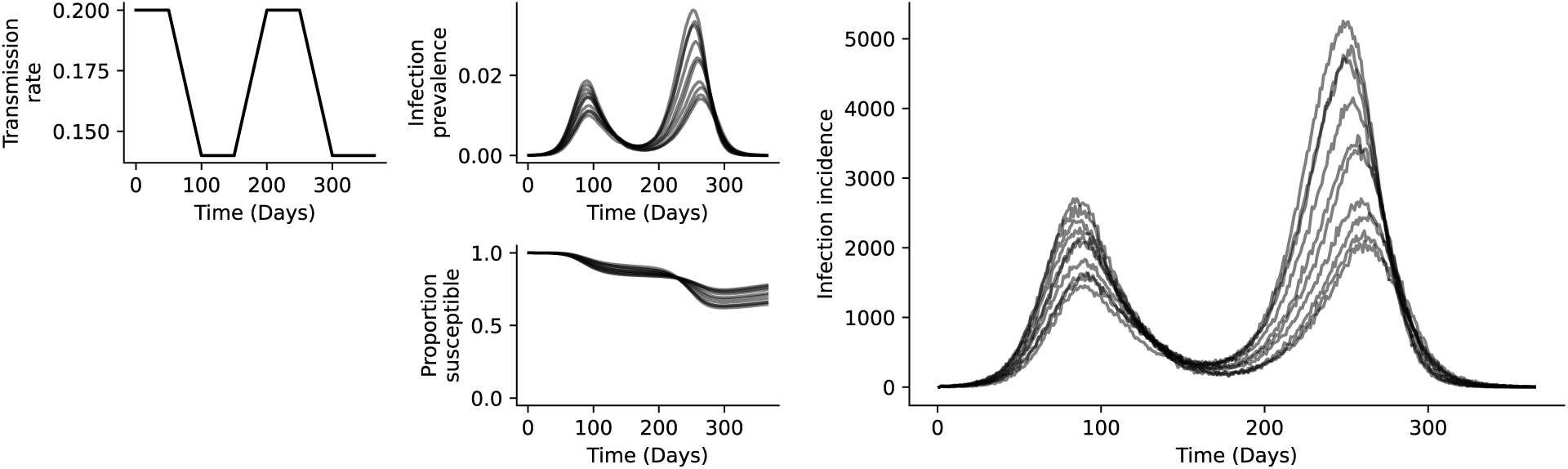
Agent-based simulation of epidemics. In the leftmost panel, we plot the unobserved time-varying transmission rate we used as input to the simulation model. In the middle two panels, we plot the unobserved infection prevalence and unobserved number of susceptibles as they vary over time, for 10 separate simulations of the model. In the right panel, we plot the unobserved true number of new infections arising each day for 10 separate simulations of the model.

Ten realizations from our model are presented in Fig. 2. Because the model is stochastic, different simulations with identical input parameter values result in different simulation outputs, as illustrated in the figure.

### 2.3 Inference for the number of infections

#### 2.3.1 Computation of the likelihood

For each survey design, we inferred the incident daily infections, denoted *i*(*t*) (i.e., we inferred *i*(1), *i*(2), *i*(3) and so forth). For computational tractability, we modelled each daily *i*(*t*) as a continuous random variable (i.e., we allow non-integer values of *i*(*t*), even though these are not possible in reality).

The same mathematical reasoning applies to both prevalence and seroprevalence surveys. The data arising from a survey round at time *t_k_* consists of the number tested positive, denoted *d_k_*, out of the total number tested, denoted *n_k_*. We assume that *d_k_* is a realization of a binomial random variable *D_k_*, whose distribution is given by:

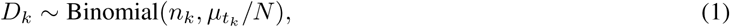

where *µ_t_k__* expresses the total number of people in the population who hold the target statuses (infectious for infection prevalence testing, and infectious or recovered for seroprevalence testing) at the time *t_k_*. We assume that *µ_t_k__* is a function of *i*(*t*), and obeys:

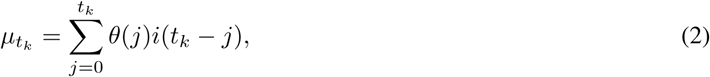

where *θ*(*j*) indicates the probability that an individual, *j* days after becoming infected, remains infectious (for infection prevalence testing), or remains either infectious or recovered (for seroprevalence testing). The values of *θ*(*j*) express population-level probabilities of being infectious or recovered (and, hence, testing positive, under our assumption of perfect test sensitivity and specificity) over time since infection rather than representing individual-level profiles of infectiousness or antibody levels through time.

We assumed that *i*(*t*) evolves in time according to a first order random walk on the logarithm scale. Specifically, we assume that:

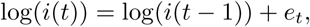

where each *e_t_*is modelled according to:

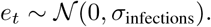

Bayesian inference to obtain the posterior distribution of *i*(*t*) conditional on simulated surveys was performed via MCMC. Full details are described in §S3.

#### 2.3.2 Computation of test positivity probabilities

Eq. (2) applies to both infection prevalence and seroprevalence surveys, although the value of *θ*(*j*) differs for each. For infection prevalence surveys, we denote by *θ*^infect^(*j*) the probability of still being infectious, and for seroprevalence surveys, we denote by *θ*^sero^(*j*) the probability of still being either infectious or recovered, *j* days after being infected. We computed each *θ*(*j*) directly from the probability distributions giving the durations that individuals spend infectious or recovered (see §2.1.1 and §S2).

We computed *θ*(*j*) under the simplifying assumption that individuals never experience multiple infections. Although this is a simplification, for our chosen epidemic configuration (§2.2), we found that only a small proportion of individuals were reinfected during the 1-year simulation period. Across the 10 replicates depicted in Fig. 2, we observed a minimum of 0.13% and a maximum of 0.20% of individuals in the population experiencing two or more infections within the yearlong simulation period, whereas between 27.6% and 40.3% of individuals in the population experienced a single infection.

### 2.4 Quantification of inference quality via CRPS

To quantify the performance of different survey designs, we evaluated the quality of surveillance data at informing *i*(*t*) using the continuous ranked probability score (CRPS) ^44^ applied to the logarithm (base *e*) transform of infection incidence data ^6^. Letting *F*_log_ *_i_*_(_*_t_*_)_ indicate the cumulative distribution function of the inferred posterior distribution of log *i*(*t*), and the true, held out value by log *ĩ*(*t*), the CRPS is given by:

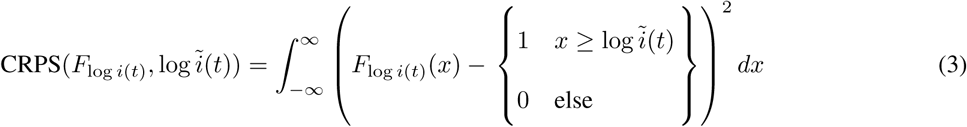

For a given data set, smaller values of the CRPS indicate a superior fit to the true values. CRPS is univariate; we obtained a single summarized score value for the entire time series of *i*(*t*) posterior distributions by summing the individual CRPS values computed according to eq. (3) over all time points, *t* = 1, 2, 3,. . ., *T*.

### 2.5 Configuration and evaluation of survey designs

#### 2.5.1 Survey designs

We considered three categories of surveillance designs: those in which infection prevalence surveys and seroprevalence surveys overlap in time and have the same sample sizes (“combined”); those in which infection prevalence and seroprevalence surveys are conducted at different frequencies and potentially overlapping (“potentially overlapping”); and those in which infection prevalence and seroprevalence tests are conducted every day throughout the epidemic (“rolling”). For combined and potentially overlapping designs, we assumed that each survey round lasted for 7 days. For each category, we considered a two-dimensional grid of possible survey design parameters; all considered designs are specified in Table 1. We assumed that the first survey (or the first day with testing, in the rolling design) occurs on day 10 of the epidemic simulation. For each surveillance design, we computed CRPS (eq. (3)) on each of 10 replicates of the epidemic scenario (§2.2). To obtain a single summary estimate of the quality of each design, we computed the median CRPS across the 10 replicates.

**Table 1:**
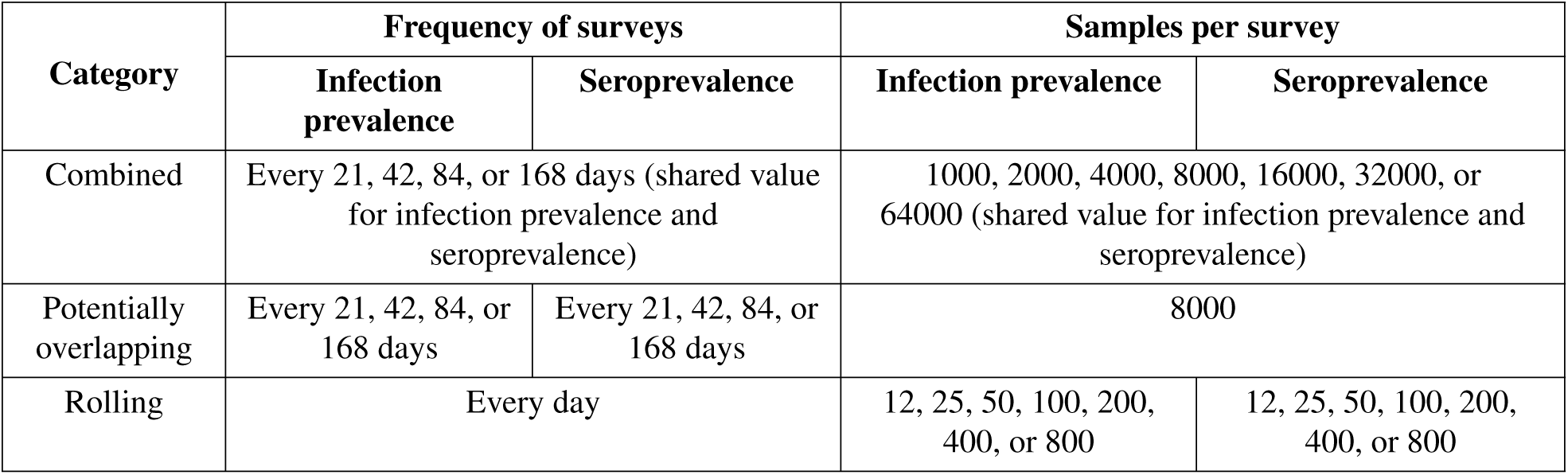
Selected configurations of survey designs.

#### 2.5.2 Misspecification of the test positivity

Given the typically long duration of seropositivity following infection, in real epidemics it can be difficult to obtain accurate estimates of *θ*^sero^(*j*), particularly during the period of emergence of a novel pathogen. Without changing the underlying simulation parameters, we performed inference under three different assumptions for the mean number of days individuals remain recovered: 300 (correct), 150, and 600; each assumption implies a *θ*^sero^ which we use for inference. In all situations, we provided the inference procedure with the correct standard deviation in the number of days individuals remain seropositive (100 days), and the correct values for *θ*^infect^(*j*).

We applied these assumptions about *θ*^sero^(*j*) to two different designs of a combined infection prevalence and seroprevalence survey: an intensive design consisting of 2000 individuals per survey type sampled every 21 days, and a sparser design consisting of 2000 individuals sampled per survey type every 84 days.

## 3 Results

### 3.1 Inferring point infection prevalence at different stages in an epidemic

To confirm that our simulation approach aligns with results from statistical theory regarding sample size for point estimates, we first investigated the impact of the sample size of an infection prevalence survey on the precision of posterior estimates of prevalence at different stages in an epidemic (i.e., different underlying true prevalence values). We configured an epidemic with a single large peak, as shown in Fig. 3a. At four selected time points, corresponding to a large range of magnitudes of the underlying true infection prevalence (up to 48% at the epidemic peak), we computed the expected posterior distribution of prevalence which would be obtained from a single infection prevalence survey at that time point, for various choices of the number of tests, i.e., the study sample size (Fig. 3b). Our simulations show diminishing returns in inference precision with increasing numbers of tests; furthermore, we observe that prevalences closer to 50% require a larger sample size in order to achieve a given precision (Fig. 3c).

**Figure 3:**
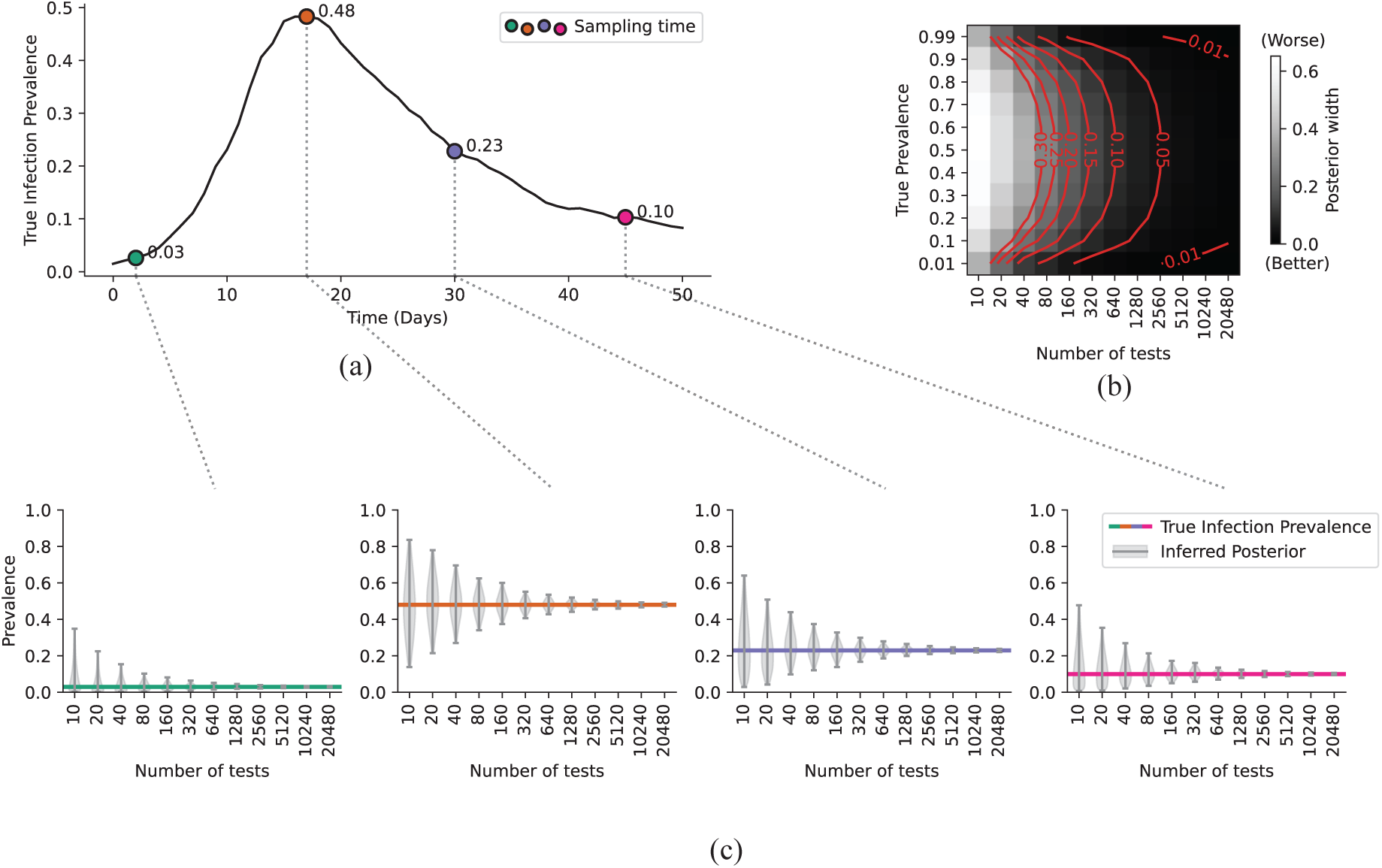
Investigating effect of numbers of tests on precision of estimates of point-in-time infection prevalence. (a) The true proportion of infected individuals over time, as simulated by one realization of our agent-based model. Four time points have been selected, representing before the peak, near the peak, shortly after the peak, and substantially after the peak. (b) The expected width of the 99% central interval of the posterior distribution as a function of the number of tests conducted and the true infection prevalence, computed on a denser grid of true infection prevalence values. Each red contour lines indicates, for a given posterior width, the number of tests required to achieve that posterior width at each value of true prevalence. (c) The expected posterior distribution of estimated infection prevalence as a function of illustrative numbers of tests conducted (i.e., sample sizes). Vertical bars indicate the bounds of the 99% central interval of the posterior distribution, while the shaded region indicates the posterior density. Horizontal lines indicate the true prevalence at each time point.

### 3.2 Investigating the impact of survey frequency and sample size on inference of infection dynamics

Across all survey configurations, we observe that the least intensive survey designs result in an inability to precisely infer infection incidence, while the most intensive survey designs lead to posterior distributions of infection incidence which are both accurate and precise (Figs. 4, 5, 6, S1). When analyzing surveys divided into discrete rounds of the same frequency and sample size (Fig. 4), we found that increasing the frequency of rounds for a given sample size, or increasing the sample size for a given survey frequency, both improved inference performance. Throughout the design space, for a given total number of tests, the lowest survey frequency that we explored (168 days) almost always led to inferior inference performance compared to more frequent survey designs with fewer participants per round (Fig. 4b). In certain regions of the design space, we found that increasing survey frequency produced greater benefit in terms of inference quality than increasing the number of participants per round. For example, survey designs with 21-day frequency tended to outperform those of similar total sample size but with 42-day frequency; and that survey designs with 42-day frequency tended to outperform those of similar total sample size but with 84-day frequency (Fig. 4b). However, the relative advantage of frequency compared to sample size becomes increasingly modest as frequency increases; furthermore, these relative advantages were limited to the subset of the design space where total tests conducted exceeded 600,000. As when inferring point prevalence (Fig. 3c), we found diminishing returns with increasing numbers of tests per round. For example, at high survey frequency (once every 21 days, shown in the leftmost column of Fig. 4a), the decrease in posterior uncertainty from 8000 to 64000 tests per survey is marginal. When we consider survey designs where infection prevalence and seroprevalence surveys share a common sample size but are conducted at different frequencies (Fig. 5), we find that either frequent infection survey rounds (with infrequent serosurvey rounds), or frequent serosurvey rounds (with infrequent infection survey rounds), can both lead to precise inference results. For “rolling” survey designs where infection prevalence and seroprevalence testing is conducted every day, superior inference performance tends to be observed with numerous infection prevalence tests and limited seroprevalence tests, compared to numerous seroprevalence tests and limited infection prevalence tests (Figs. 6, S1).

**Figure 4:**
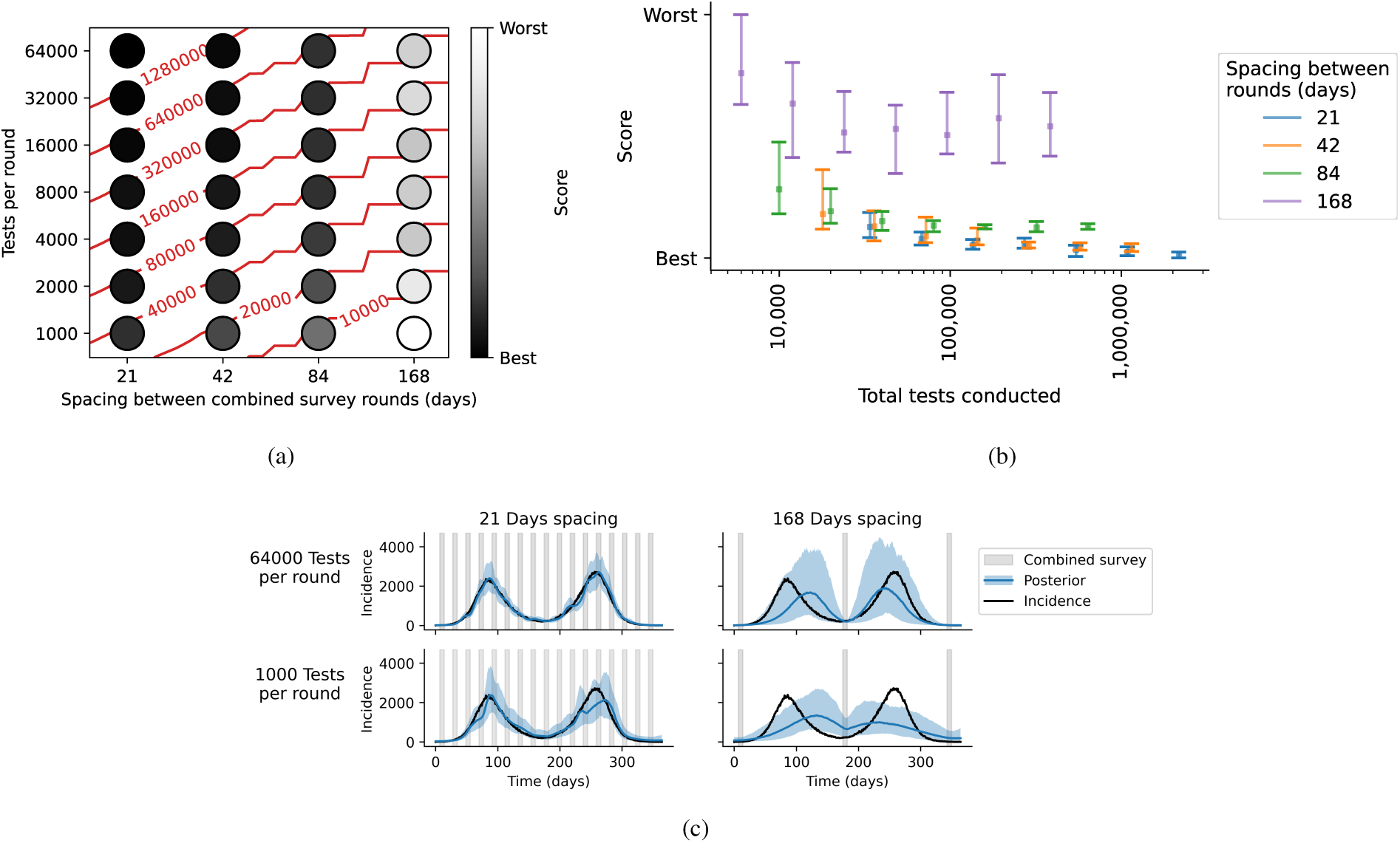
Evaluating inference performance when infection prevalence and seroprevalence surveys overlap (combined survey designs). (a) For each combined survey design, consisting of the stated survey frequency and tests per round (where rounds of seroprevalence and infection prevalence surveys are conducted at at overlapping times), the median value of the CRPS score averaged over 10 replicates. Red contour lines represent the total number of tests conducted across both survey types. (b) For each combined survey design, the median (dot) and range (bar) of scores for the 10 replicates at each spacing, plotted at the corresponding total tests conducted across both survey types throughout the epidemic. (c) For four selected survey designs, including the most intensive and least intensive designs (in terms of total numbers of tests) within the space of possible designs considered in this experiment, the posterior distribution for the inferred incident infections each day overlaid on the true, unobserved number of incident infections each day. The shaded curve indicates the 99% central interval of the posterior distribution, the blue line indicates the posterior median, and the black line indicates the true incident infections. Shaded boxes indicate periods of time when survey data were collected.

**Figure 5:**
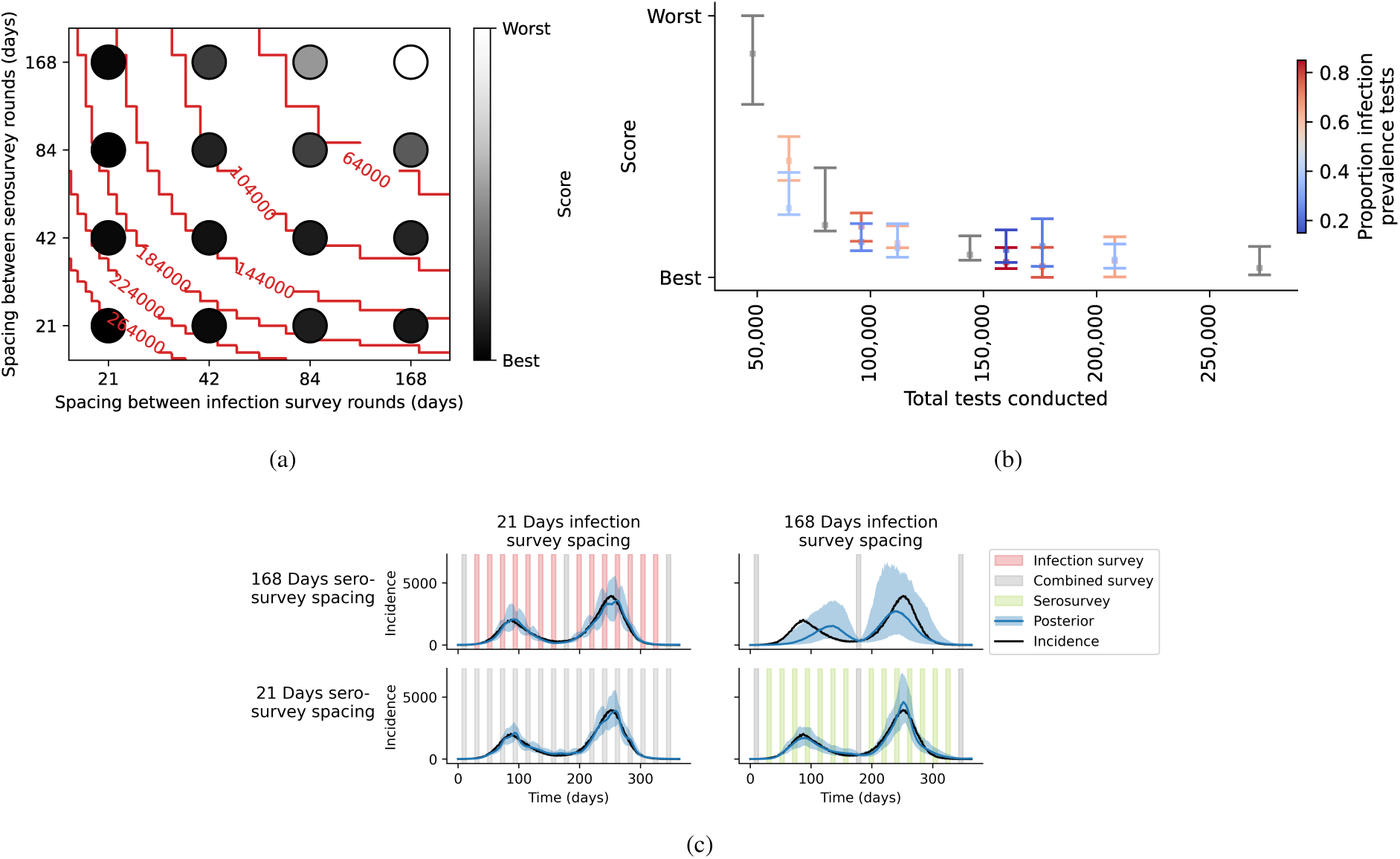
Evaluating inference performance when infection prevalence and seroprevalence surveys differ in frequency (potentially overlapping survey designs). (a) For each potentially overlapping survey design, consisting of 8000 tests per round of surveying (where rounds of seroprevalence and infection prevalence surveys are conducted at potentially different frequencies, and may or may not overlap), the median value of the CRPS score averaged over 10 replicates. Red contour lines represent the total number of tests conducted across both survey types. (b) For each survey design, the median (dot) and range (bar) of scores for the 10 replicates, plotted at the corresponding total tests conducted across both survey types throughout the epidemic. The color indicates the proportion of all tests conducted which were infection prevalence rather than seroprevalence (e.g., the design with infection prevalence surveys conducted at 168 day spacing and serosurveys conducted at 84 day spacing, is colored according to 0.375 of tests being infection prevalence). (c) For four selected survey designs, including the most intensive and least intensive designs (in terms of total numbers of tests) within the space of possible designs considered in this experiment, the posterior distribution for the inferred incident infections each day overlaid on the true, unobserved number of incident infections each day. The shaded curve indicates the 99% central interval of the posterior distribution, the blue line indicates the posterior median, and the black line indicates the true incident infections. Shaded boxes indicate periods of time when survey data were collected.

**Figure 6:**
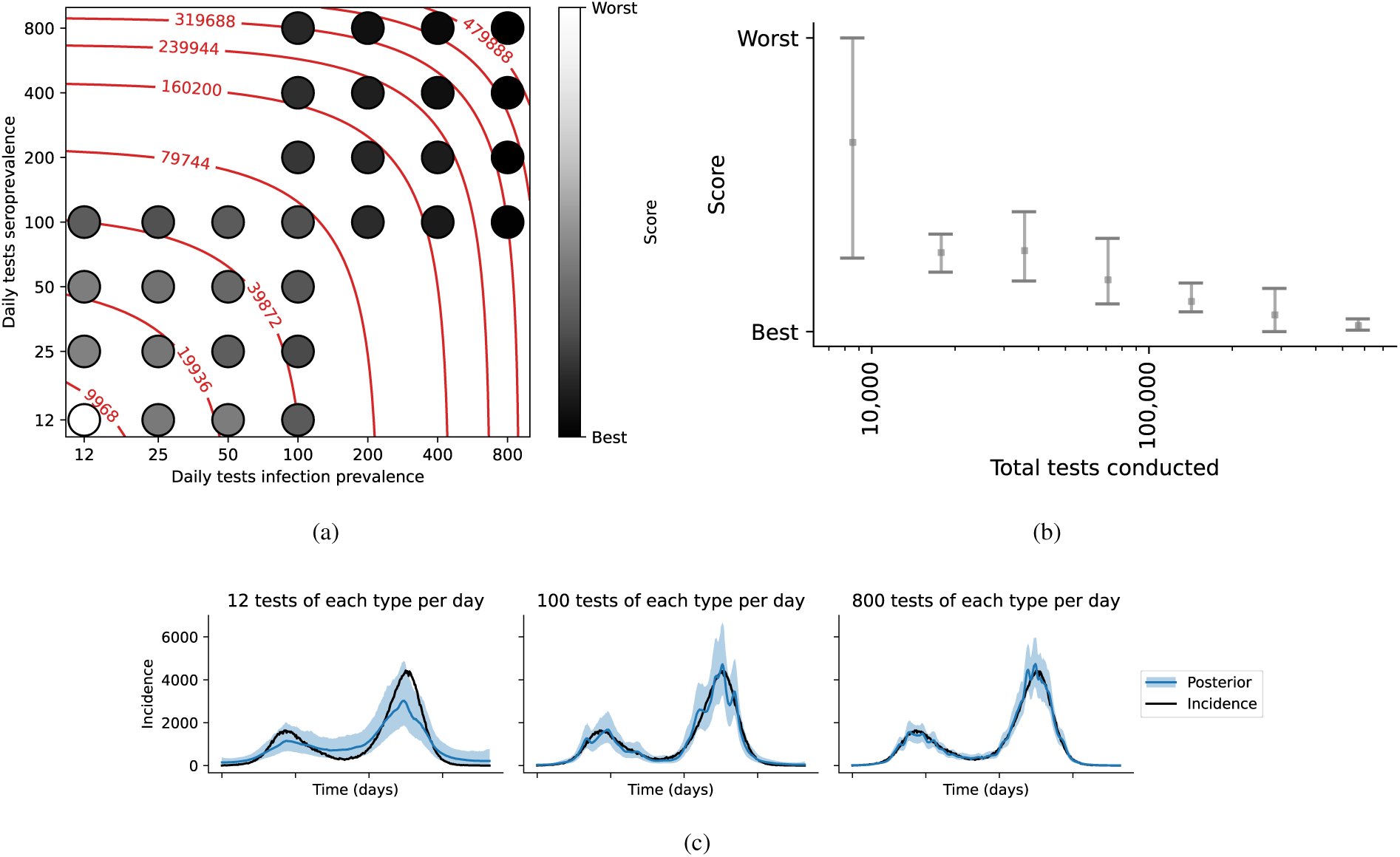
Evaluating inference performance when infection prevalence and seroprevalence testing occurs every day (rolling survey designs). (a) For each rolling survey design, consisting of the number of tests per day of surveying (where each day uses both seroprevalence and infection prevalence testing), the median value of the CRPS score averaged over 10 replicates. Red contour lines represent the total number of tests conducted across both survey types. (b) For each survey design in which infection prevalence and seroprevalence testing have the same sample size, the median (dot) and range (bar) of scores for the 10 replicates, plotted at the corresponding total tests conducted across both survey types throughout the epidemic (ranging from 12 tests per day of each type, to 800 tests per day of each type). (c) For three selected survey designs, including the most intensive and least intensive designs (in terms of total numbers of tests) within the space of possible designs considered in this experiment, the posterior distribution for the inferred incident infections each day overlaid on the true, unobserved number of incident infections each day. The shaded curve indicates the 99% central interval of the posterior distribution, the blue line indicates the posterior median, and the black line indicates the true incident infections.

In §S4 we present additional results when infection prevalence and seroprevalence tests have imperfect sensitivity and specificity, from both a simulation-based analysis (as above) and a distinct theoretical analysis. When considering single point-in-time estimates of prevalence (as in Fig. 3) via our theoretical analysis, we observe that imperfect testing conveys less information than perfect testing, although the magnitude of this effect varies sharply with prevalence. As prevalence approaches zero, the number of imperfect tests that would be required to achieve the same information as perfect testing rapidly increases. In our time series inference problems for infection incidence (simulation-based), because information about incidence is shared across time, the degradation in overall inference performance (as assessed by CRPS) under rolling designs due to imperfect testing is generally minor (Fig. S4). And, under a rolling design, a modest increase in sample size allows similar inference performance to be achieved despite imperfect sensitivity and specificity.

### 3.3 Inaccurate knowledge of the timing of seroreversion can degrade inference quality

Misspecification of *θ*^sero^(*j*) is observed to lead to biased posterior distributions of infection incidence, with *i*(*t*) consistently overestimated when the duration of seropositivity is underestimated, and *i*(*t*) occasionally underestimated when the duration of seropositivity is overestimated (Fig. 7). In the early phase of the epidemic (e.g., first 100 days), bias is limited and estimates of *i*(*t*) are relatively accurate. This is because all infections are recent and the assumed and true distributions of *θ*^sero^(*j*) have not yet diverged, particularly when the decay rate is assumed to be slower than the true distribution (assumed and actual distributions are approximately the same for the first 100 days since infection; Fig. 7a). As the epidemic progresses, individuals that were infected sufficiently far in the past (such that the assumed and true distributions of *θ*^sero^(*j*) have diverged) will accumulate and increasingly impact the inference of *i*(*t*). This result also highlights that the information provided about infections from each survey type changes over the course of the epidemic. For example, fewer individuals will be seropositive in the earlier phases of an epidemic and so seroprevalence surveys provide less information relative to later in the epidemic. Finally, we observe that when *θ*^sero^(*j*) is misspecified, increasing the survey frequency (with the same number of individuals per survey) can lead to increasingly inaccurate and overconfident estimates of *i*(*t*) (Fig. 7b, left column).

**Figure 7:**
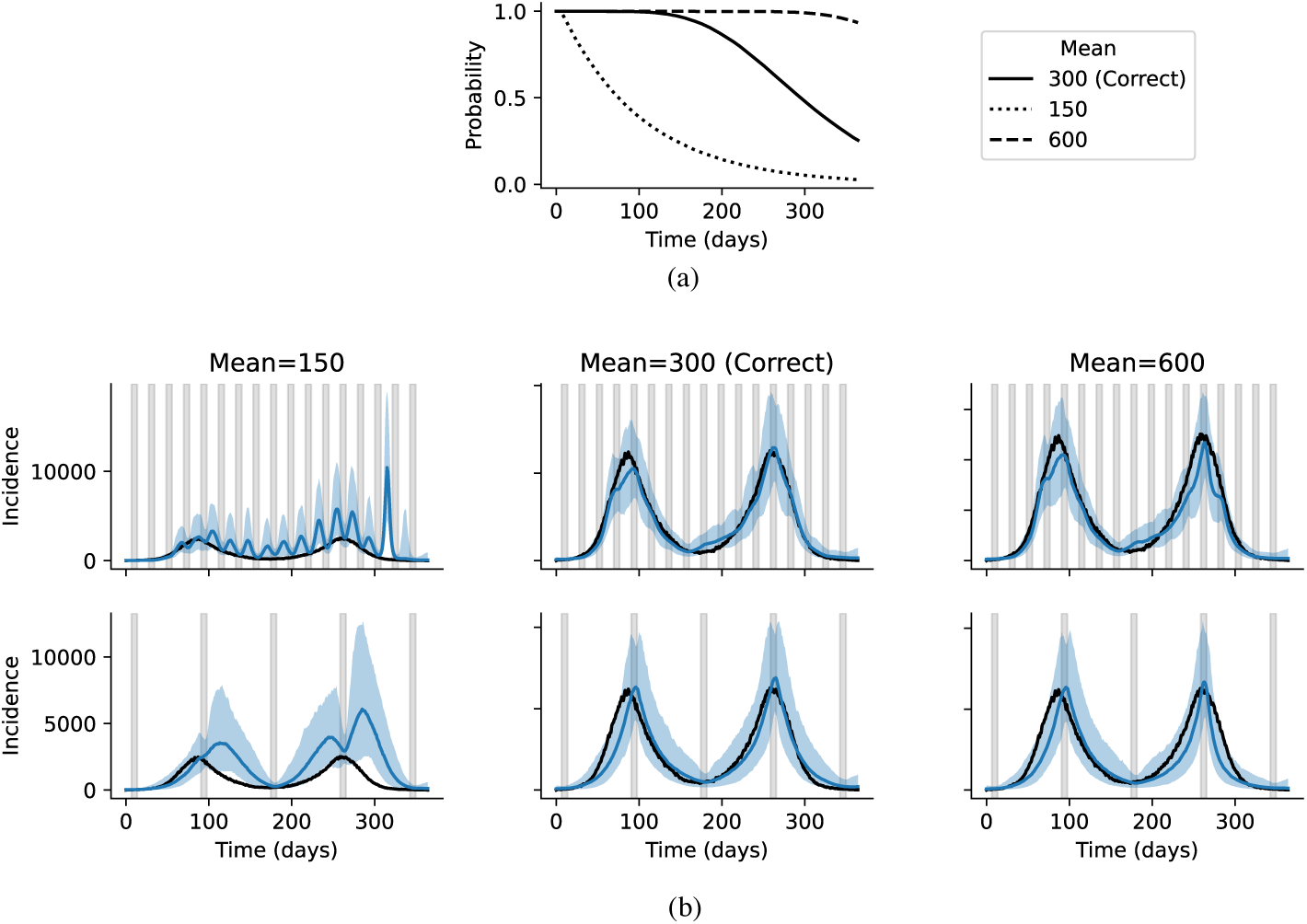
Effect of misspecification of serology test positivity probabilities on inference results for the true number of infections. (a) Three selected assumptions for the probability of being positive for antibodies as a function of time since infection, labelled by the corresponding mean number of days individuals would test positive for antibodies after ceasing to be infectious under that assumption. (b) Posterior distributions for the true number of infections resulting from each assumption about test positivity, in a setting of combined infection prevalence and seroprevalence surveys with intensive frequency (top row) and sparse frequency (bottom row). The shaded curve indicates the 99% central interval of the posterior distribution, the blue line indicates the posterior median, and the black line indicates the true incident infections. Shaded boxes indicate periods of time when combined survey data were collected.

## 4 Discussion

Our study has introduced a framework for using epidemiological modelling to inform infection survey design. Using our framework to investigate a multitude of survey designs, we found that when infection prevalence and seroprevalence testing is conducted in combined discrete survey rounds (21, 42, 84, or 168 days between rounds), higher survey frequency typically produced greater benefit than a higher number of tests per round, for a given total sample size. Irrespective of the number of tests per round, the worst performance was observed when surveys were insufficiently frequent (168 days in our study). Rolling surveys exhibited superior performance on average across all designs explored in our analysis. They also displayed lower variance across replicates, and as such were never found to be the worst design for any given total sample size, supporting their deployment in real world studies (e.g., as used to investigate COVID-19 prevalence and seroprevalence in England ^32^).

We anticipate that timing parameters of the pathogen (e.g., generation interval) are likely to be important factors for optimal survey design, and it would be productive to apply our framework to different epidemiological contexts. The surveillance designs we identified as most efficient in this paper are contingent on the epidemic scenario that we analyzed, and our results will not necessarily generalize to pathogens which differ substantially from the one we simulated. For example, we expect that in general more frequent survey rounds would be required to capture the underlying infection dynamics of a pathogen with a larger basic reproduction number or shorter generation interval, but this would also depend on other factors such as intervention measures which are not known in advance of deploying surveys.

Consistent with statistical theory ^38^, both our point estimates and time series estimates demonstrated diminishing returns in inference quality as total sample size increased. In our study, sample sizes greater than approximately 10,000 per month were observed as being increasingly inefficient. Infection prevalence and seroprevalence surveys conducted for COVID-19 in the United Kingdom (REACT and CIS) involved sample sizes of hundreds of thousands of individuals tested per month ^32;36^, significantly exceeding the sample sizes investigated in our study. However, these real-life studies were designed to have sufficient power to capture information about COVID-19 prevalence and seroprevalence within specific age groups and geographical regions, as well as to investigate other risk factors or changes over time ^15^; these objectives, and the age, geographical, or other heterogeneities underlying them, have not been considered in our study. Instead, our results are relevant to population-level estimates of infection incidence through time (or alternatively to estimates of the infection incidence in a given cohort within a larger population). However, it would be straightforward to adapt our framework to include population heterogeneity such as demographic or spatial structure; our approach could then be used to evaluate survey designs according to more complex objective functions incorporating the considerations which have guided the design of real infection surveys such as REACT and CIS. Likewise, adaptations of the analysis could be used to evaluate the value of more complex sampling designs, such as convenience samples from healthcare settings. Additionally, it would be possible to extend our framework to investigate the impact of, for example, vaccination ^1^.

We found that infection prevalence samples typically provided more information to estimate infection dynamics than seroprevalence surveys. Amongst rolling designs, those with a majority of infection prevalence testing tended to slightly outperform those with a majority of seroprevalence testing (Fig. S1). This finding is not surprising given that the objective function we used to evaluate survey quality, which assessed the accuracy and precision of inference of true infections arising through time. Seroprevalence surveys are typically used to evaluate point-in-time history of exposure in a population, a distinct objective to that used in our study. Furthermore, the relative value of infection prevalance and seroprevalence surveys will depend on the duration of test positivity of each test type.

The possibility of combining information from infection prevalence surveys with information from cases to obtain more precise time-varying estimates of prevalence has been investigated ^29^. Our framework for evaluating survey designs could be extended to include simulations of cases, and the incorporation of case data when inferring the true infection incidence. Such a procedure is expected to have benefits but also potential drawbacks. It could lead to more precise estimates of true infection incidence at a given level of survey intensity due to additional information present in the cases, particularly during periods between surveys; this could be particularly helpful for improving the performance of surveillance designs where surveys are conducted infrequently. However, using case data may lead to biases when case ascertainment varies significantly over time, and it presents computational challenges and may be inappropriate in settings where cases data are of low quality or unreliable ^28;2^.

In our study, we considered survey design parameters from pre-selected grids of candidate values; such an approach is unlikely to identify the exact survey design parameters which maximize the objective function. Furthermore, we restricted our analysis to two-dimensional grids of design parameter values; we did not investigate the impact of factors such as the timing of the initial survey round. Later timing of the initial survey round may enable superior performance for survey designs with long gaps between survey rounds (i.e., the 168-day spacing in Fig. 4c); however, in actual practice, it would be difficult to determine optimal timing of an initial survey in advance of knowing how the epidemic will progress. It would be productive to integrate our framework for simulating and evaluating design surveillance strategies with a formal optimization scheme which proposes promising candidates designs, thus exploring (potentially high dimensional) spaces of potential surveillance designs and identifying the optimal design subject to desired constraints on resources availability. We anticipate methods from Bayesian optimal design ^39;35^, or Bayesian optimization ^17^ to be useful for this purpose. Likewise, our model-based approach could be used to conduct a power analysis identifying the number of tests required to infer infection incidence (or other related quantities) at a given time with a given margin of error.

Our investigation of misspecification of the duration of seropositivity revealed that inaccurate knowledge of this quantity substantially impeded the ability to accurately infer infections from seroprevalence survey data (in particular, a significant erroneous oscillatory pattern was inferred in infections when the duration of seropositivity was underestimated). Our results are in accordance with previous findings that failing to account for seroreversion caused underestimation of infection prevalence from serosurveys for SARS-CoV-2 in England ^10^. Further work to obtain better approaches to estimating the duration of seropositivity would be valuable. In order to leverage information about the duration of seropositivity which is available in the population-level seroprevalence revealed by the seroprevalence surveys, it would be productive to consider a joint inference procedure for estimating the duration of seroprevalence and incident infections.

Our investigation evaluated survey designs based solely on their ability to inform the historical counts of new infections, *i*(*t*). Although knowledge of the underlying infection burden is an intrinsically useful epidemiological quantity, the information on past infections inferred from the data collected is also a crucial input for prediction tasks. Key examples include forecasts of future epidemic activity or the impact of interventions, or future infection incidence within finer age-specific or location-specific groups, or forecasting of hospitalisations and deaths (all of which rely on understanding of infection trends by age-group and severity). More sophisticated objective functions could be derived which evaluate the quality of survey designs at informing these downstream quantities of interest, enabling our framework to help identify the best survey designs for a variety of public health tasks.

## Statements and Declarations

### Data transparency

All software and code notebooks to reproduce the analyses in this paper are available open source at https://github.com/epi-analytics-surveillance/designing-surveys.

