## Supplemental information for "Designing infection prevalence and seroprevalence surveys for learning infection dynamics"

### S1 Supplementary results

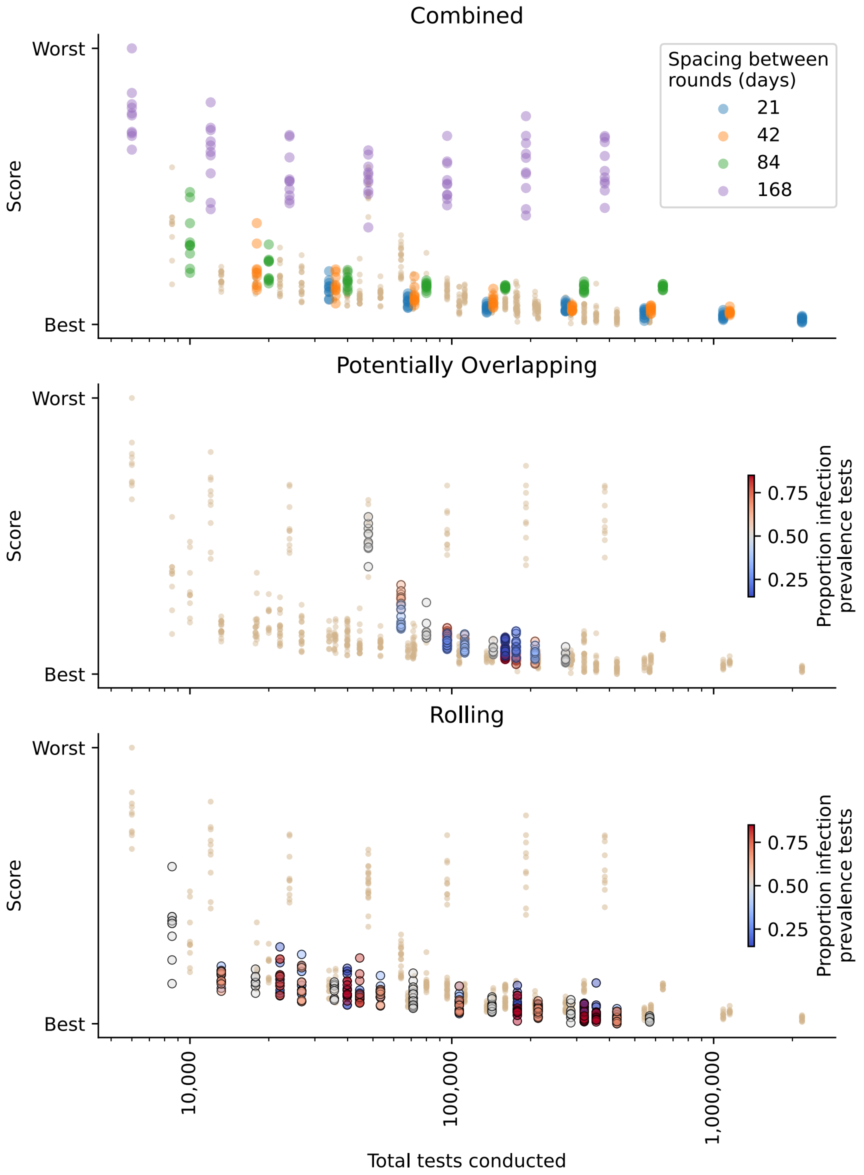


Figure S1: **Evaluating inference performance for all surveillance designs.** For each survey design, the CRPS scores for the 10 replicates at each spacing and corresponding total tests conducted throughout the epidemic. In each panel, the results for the category of survey design (see Table 1) indicated by the panel title are shown in the colors indicated by each panel legend, while all results corresponding to the other two categories of designs are shown in brown.

#### S2 Epidemic simulation details

The model involved $j = 1, 2, 3, \ldots, N$ agents, each representing an individual person in the population. A discrete set of time steps was used, labeled $t = 0, 1, 2, 3, \ldots, T$. The duration between consecutive time steps is denoted $\Delta t$; throughout this paper, we assumed that $\Delta t = 1$ day.

At each time, each agent has one of three possible statuses: susceptible, infectious, and recovered. At time $t$, the total number of susceptible individuals is denoted $S(t)$, the total number of infectious individuals is denoted $I(t)$, and the total number of recovered individuals is denoted $R(t)$. Individuals classified as recovered are assumed to be immune to re-infection.

To simulate transmission of the pathogen between time $t$ and time $t+1$, each individual $j$ who was infectious at time $t$ randomly selects some number $n_j^{\text{transmit}}$ of susceptible individuals (to whom the infection will be transmitted) according to:

$$n_j^{\text{transmit}} \sim \text{Poisson}(\rho(t)S(t)/N).$$

where $\rho(t)$ indicates the transmission rate at time $t$. If $n_j^{\text{transmit}} > S(t)$, it is set as $n_j^{\text{transmit}} = S(t)$. Then $n_j^{\text{transmit}}$ randomly selected susceptible individuals become infectious, and $S(t)$ is immediately updated according to $S(t) \leftarrow S(t) - n_j^{\text{transmit}}$. This process is repeated for each individual who was infectious at time $t$.

We simulated progression of individuals from infectious to recovered, and recovered individuals back to susceptible, by drawing the number of days each individual retained their infectious or recovered status according to discrete probability distributions. Specifically, at the time when an individual $j$ acquires infectious or recovered status, the number of days they remain with that status is denoted $n_j^u$, and is sampled according to:

$$n_j^u \sim f^{\text{u}},$$

for $u \in \{\text{infectious}, \text{recovered}\}$. After $n_j^u$ days have passed, the individual $j$ is moved to recovered if they were infectious, and back to susceptible if they were recovered. $f^{\text{infectious}}$ and $f^{\text{recovered}}$ correspond to discrete probability mass functions which are specified in order to represent the pathogen of interest. For our study, we assumed discretized gamma distributions (specified by the mean $\mu$ and standard deviation $\sigma$). We assumed $\mu^{\text{infectious}} = 7$, $\sigma^{\text{infectious}} = 2$, $\mu^{\text{recovered}} = 300$, and $\sigma^{\text{recovered}} = 100$.

The model was initialized by assuming that at the initial time $t = 0$, some initial number of infectious individuals, denoted $I(t = 0)$, are infectious, while the remaining $N - I(t = 0)$ individuals are susceptible. The population size $N$ was fixed (by construction).

To obtain the true series of new infections, we counted the number of individuals acquiring infectious status at each time $t$. We denoted the ground truth number of new infections at time $t$ by $\tilde{i}(t)$. Due to our choice of $\Delta t = 1$ day,

$\tilde{i}(t)$ represents the number of daily new infections.

#### S3 Inference details

For each inference problem (see §2.3.1), we ran two chains of the No-U-Turn (NUTS) sampler in Stan[8]. We tuned the adaptation target Metropolis acceptance rate (`adapt_delta`) to 0.99, and the maximum depth of NUTS trees (`max_treedepth`) to 20.

We assessed convergence by computing the Gelman $\hat{R}$ statistic[19] for each value of $\log(i(t))$, i.e., for $\log(i(1))$, $\log(i(2))$, and so forth. At first, we ran each chain for a total of 1200 iterations, with the first 400 discarded as burnin. If all $\hat{R}$ values achieved $\hat{R} < 1.05$, we treated the chains as converged and used their samples for downstream analyses. If any $\hat{R}$ value exceeded $1.05$, we doubled the number of total MCMC iterations and repeated the MCMC inference (including further doubling as needed) until achieving all $\hat{R} < 1.05$.

We found that when MCMC chains were initialized by sampling from the prior distribution, or in any other location which is not near to the posterior mode, our MCMC approach as outlined above is likely to fail to converge, particularly for surveillance designs with larger spacing between rounds of surveys. We thus developed a method to initialize MCMC at plausible values without providing direct access to the true values $i(t)$. For each epidemic, we simulated a time series of case data, denoted $c(t)$, by assuming that each newly infected individual is reported as a case with probability $\phi$. (Thus, with probability $1 - \phi$, an infected individual is never reported as a case and does not appear in $c(t)$.) For simplicity, we neglected delays between infection onset and case reporting and assumed that cases were reported on the same day they became infected. For our analysis in this paper, we set $\phi = 0.5$. To compute MCMC initializations, we assumed access to $c(t)$, but not to the fact that $c(t)$ was generated using $\phi = 0.5$. We generated candidate MCMC initializations of $i(t)$ by drawing a candidate value $\phi'$ from $\text{Uniform}(0.2, 0.8)$, and setting $i(t) = c(t)/\phi'$. We repeated this procedure 25 times, and selected the two candidate initializations achieving the highest log probability under our posterior as the initializations for the two MCMC chains.

We assumed that the random walk prior standard deviation parameter, $\sigma_{\text{infections}}$, itself obeyed a truncated normal prior with mean zero, standard deviation $\sigma_{\text{hyper}}$, truncated below at zero, and with no upper truncation:

$$\sigma_{\text{infections}} \sim \mathcal{N}^+(0, \sigma_{\text{hyper}}).$$

Each inference problem was assumed to have its own value of $\sigma_{\text{infections}}$, i.e., different surveillance designs were allowed to take different values of the parameter. However, we assumed a common, constant value of $\sigma_{\text{hyper}}$ for all inferences in this paper.

To inform appropriate values of $\sigma_{\text{hyper}}$, we took one of the stochastic epidemic simulations (§2.1.1) and computed

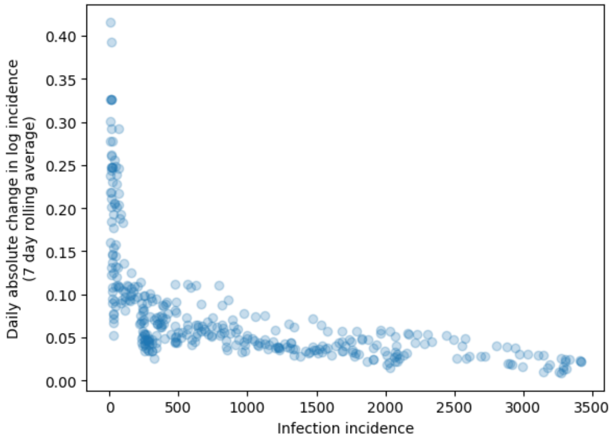


Figure S2: The 7-day rolling average in daily absolute change in log incidence as a function of the value of infection incidence, computed from one of the stochastic realizations of the epidemic scenario §2.1.1. Each dot corresponds to one time point within the epidemic, with its x-axis value being the value of infection incidence at that time point, and its y-axis value being the 7-day rolling average in daily absolute change in log incidence.

the daily absolute changes in log incidence, plotted in Fig. S2. At the beginning and end of the epidemic, infection incidence is close to zero, and small daily changes in infection incidence correspond to large log differences; however, for the rest of the epidemic, typical values of the daily change in log incidence are close to 0.05. We thus set $\sigma_{\text{hyper}} = 0.05$.

To evaluate the relationship between the inferred $\sigma_{\text{infections}}$ and surveillance design, we plotted the posterior of $\sigma_{\text{infections}}$ for each surveillance design from Fig. 4. These results are shown in Fig. S3, and suggest that values of $\sigma_{\text{infections}} \approx 0.15$ are inferred for the majority of surveillance designs.

### S4 Imperfect test sensitivity and specificity

In this section, we will relax the assumption of perfect test accuracy and consider tests whose results do not always perfectly reflect the tested individual's status. We will consider both imperfect sensitivity (e.g., for infection prevalence testing, an individual who is truly infectious may nevertheless test negative with some probability) and imperfect specificity (e.g., for infection prevalence testing, an individual who is truly not infectious may nevertheless test positive with some probability). We will assume that the values of sensitivity and specificity for each test type (infection prevalence and seroprevalence) are known (i.e., we will not misspecify sensitivity or specificity, nor attempt to infer them from data).

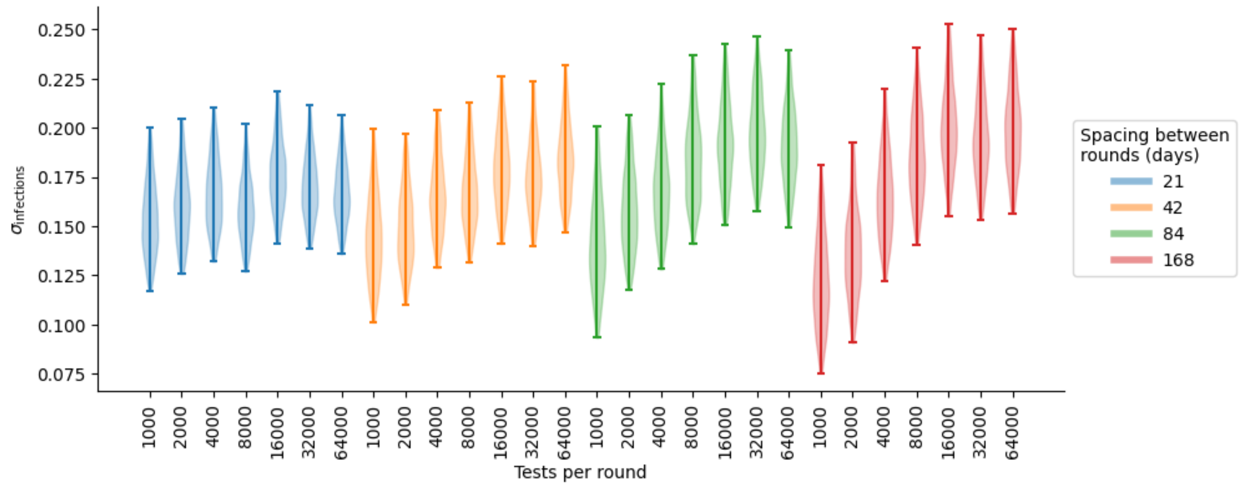


Figure S3: Inferred posterior distribution for $\sigma_{\text{infections}}$, for each of the overlapping survey designs as analyzed in Fig. 4.

#### S4.1 Likelihood for infection incidence under imperfect testing

The same equations apply for both infection prevalence and seroprevalence. Let $\pi_{\text{actual}}(t)$ denote the true prevalence (or seroprevalence) expressed as a proportion of the population, i.e., $\pi_{\text{actual}}(t_k) = \mu(t_k)/N$ (see eq. (2)). When an imperfect test with sensitivity $S$ and specificity $C$ is conducted, we denote the corresponding probability of testing positive by $\pi_{\text{obs}}(t)$, which obeys:

$$\pi_{\text{obs}}(t) = S \times \pi_{\text{actual}}(t) + (1 - C)(1 - \pi_{\text{actual}}(t)). \tag{4}$$

To compute the likelihood under imperfect testing, we thus modify eq. (1):

$$D_k \sim \text{Binomial}(n_k, \pi_{\text{obs}}(t_k)). \tag{5}$$

In this likelihood, the unknown parameter vector (infection incidence, $i(t)$) enters eq. (5) via $\pi_{\text{obs}}(t_k)$, which depends on $\pi_{\text{actual}}(t_k)$ according to eq. (4); $\pi_{\text{actual}}(t_k)$ is itself a function of $\mu(t_k)$, and $\mu(t_k)$ is a function of $i(t)$ (see eq. (2)). The simulation and inference procedure for analyzing survey designs with imperfect testing is unchanged aside from this modification to the likelihood.

We set $S = 0.9$ and $C = 0.99$, for both infection prevalence testing and seroprevalence testing, and evaluated the inference performance for 10 replicates of rolling designs (see Table 1). The results are plotted alongside the original results with $S = 1$ and $C = 1$ in Fig. S4. We observe that imperfect testing marginally worsens the CRPS scores at any given sample size; however, considering the substantial variation from replicate to replicate, there is a overlap between the results with imperfect testing and the results with perfect testing.

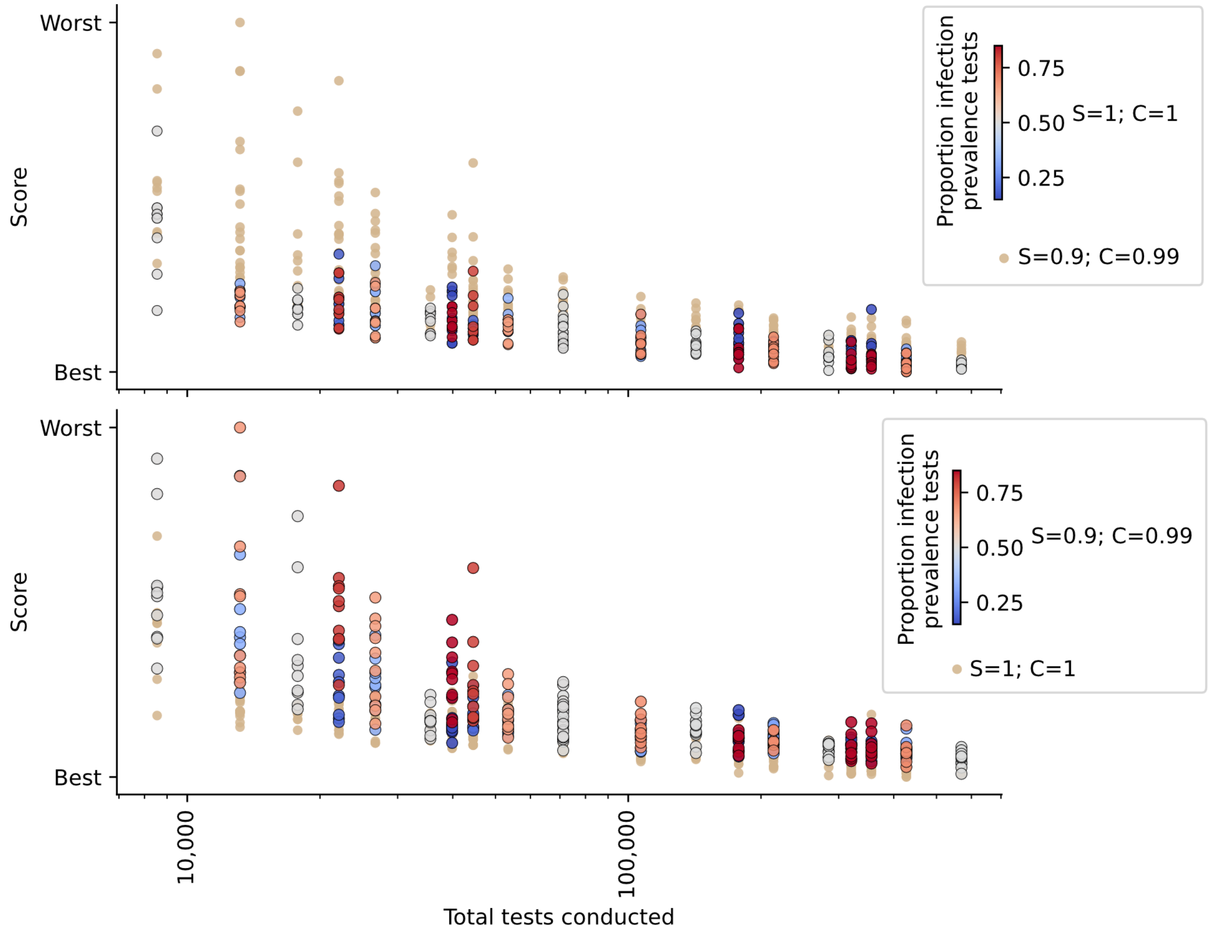


Figure S4: **Evaluating inference performance for rolling survey designs with potentially imperfect testing.** For each sample size, the CRPS scores for the 10 replicates at each spacing and corresponding total tests conducted throughout the epidemic are plotted as dots. Both panels use rolling survey designs (see Table 1) and contain the same data points. In the top panel, results based on perfect testing ($S = 1$; $C = 1$) are indicated in red, grey, and blue while results based on imperfect testing ($S = 0.9$; $C = 0.99$) are indicated in brown. In the bottom panel, results based on perfect testing ($S = 1$; $C = 1$) are indicated in brown while results based on imperfect testing ($S = 0.9$; $C = 0.99$) are indicated in red, grey, and blue.

We illustrate the relationship between imperfect test accuracy and inference performance by plotting three representative inferred $i(t)$ trajectories in Fig. S5. We plot results for 25 perfect tests per day, 25 imperfect tests per day, and 50 imperfect tests per day (all with the same underlying epidemic). The $i(t)$ inferred with 25 imperfect tests per day is visibly inferior to that inferred with 25 perfect tests per day, with larger uncertainty (particular when incidence is low) and lower accuracy. However, with 50 imperfect tests per day, the inferred $i(t)$ trajectory is visibly similar to that inferred with 25 perfect tests per day.

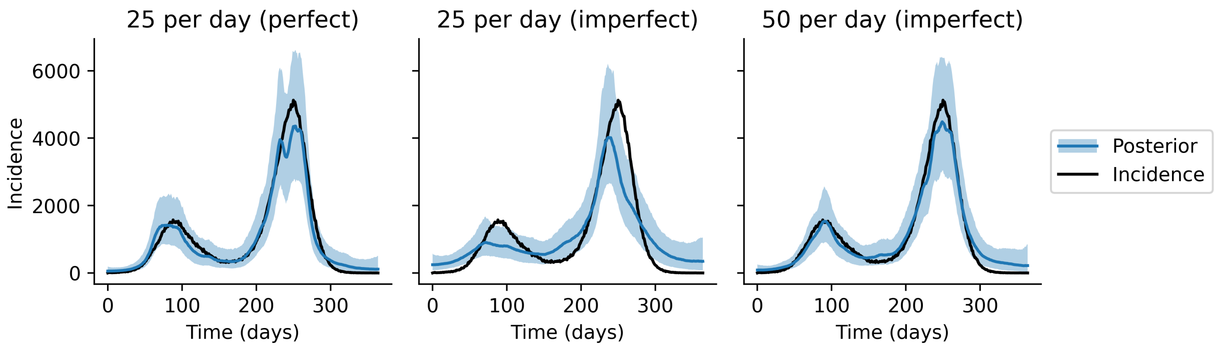


Figure S5: **Example inference performance for rolling survey designs with potentially imperfect testing.** For 25 tests per day of each type with perfect sensitivity and specificity (left), 25 tests per day of each type with $S = 0.9$ and $c = 0.99$ (middle), and 50 tests per day of each type with $S = 0.9$ and $C = 0.99$ (right), we plot the posterior distribution for the inferred incident infections each day overlaid on the true, unobserved number of incident infections each day, for one epidemic simulation. The shaded curve indicates the 99% central interval of the posterior distribution, the blue line indicates the posterior median, and the black line indicates the true incident infections.

#### S4.2 Theoretical analysis

Eq. (5) is amenable to analysis based on the Fisher information (denoted $\mathcal{I}$). Given a random variable whose distribution depends on parameters whose values are unknown, Fisher information quantifies the extent to which observations of the random variable inform the value of the unknown parameters. For a single unknown parameter, $1/\mathcal{I}$ provides a lower bound on the variance of an unbiased estimator of the parameter; thus, maximizing $\mathcal{I}$ can be seen as equivalent to minimizing uncertainty of parameter estimates.

In our analysis, for simplicity, we consider estimation of the prevalence (or seroprevalence) at a single point in time and neglect computation of the incidence. This analysis differs from that conducted in the main text or §S4.1, as it does not incorporate any prior distribution, does not compute incidence, and does not use CRPS as the objective function.

Our unknown parameter is the actual prevalence (or seroprevalence) as a proportion of the population, denoted $\pi_{\text{actual}}$; the data consists of the number tested positive (modelled with the random variable $D$) upon conducting $n$ potentially imperfect tests with sensitivity $S$ and specificity $C$. The likelihood in this setting is:

$$D \sim \text{Binomial}(n, \pi_{\text{obs}}),$$

where,

$$\pi_{\text{obs}} = S\pi_{\text{actual}} + (1 - C)(1 - \pi_{\text{actual}}).$$

The Fisher information for the binomial distribution is given by:

$$\mathcal{I}(\pi_{\text{obs}}) = \frac{n}{\pi_{\text{obs}}(1-\pi_{\text{obs}})}.$$

Applying the change of variables from $\pi_{\text{obs}}$ to $\pi_{\text{actual}}$, we obtain:

$$\begin{aligned}\mathcal{I}(\pi_{\text{actual}}) &= \mathcal{I}(\pi_{\text{obs}})\left(\frac{d\pi_{\text{obs}}}{d\pi_{\text{actual}}}\right)^2 \\ &= \frac{nJ^2}{(1+J\pi_{\text{actual}}-C)(C-J\pi_{\text{actual}})},\end{aligned}$$

where we define $J = S + C - 1$.

Under perfect testing, we have $S = C = 1$, and thus:

$$\mathcal{I}^{\text{perfect}}(\pi_{\text{actual}}) = \frac{n^{\text{perfect}}}{\pi_{\text{actual}}(1-\pi_{\text{actual}})}.$$

We compute the ratio between the Fisher information with $n$ imperfect tests, and the Fisher information with $n^{\text{perfect}}$ perfect tests:

$$\frac{\mathcal{I}}{\mathcal{I}^{\text{perfect}}} = \frac{n}{n^{\text{perfect}}} \times \frac{J^2\pi_{\text{actual}}(1-\pi_{\text{actual}})}{(1+J\pi_{\text{actual}}-C)(C-J\pi_{\text{actual}})}.$$

On the basis of this ratio, we can compute the sample size $n$, under imperfect testing, that achieves equivalent Fisher information to perfect testing with sample size $n^{\text{perfect}}$:

$$n = \frac{n^{\text{perfect}}(1+J\pi_{\text{actual}}-C)(C-J\pi_{\text{actual}})}{J^2\pi_{\text{actual}}(1-\pi_{\text{actual}})}. \tag{6}$$

Eq. (6) is visualized in Fig. S6, for various selected values of $S$ and $C$, over a range of prevalences typical of the infection prevalence occurring in our epidemic simulations in this paper (see Fig. 2). We observe that imperfect testing conveys less information than perfect testing, although the magnitude of this effect varies sharply with prevalence. As prevalence approaches zero, the number of imperfect tests that would be required to achieve the same information as perfect testing rapidly increases. However, we note that Fig. 2 merely analyzes a single point-in-time estimate of prevalence. In our time series inference problems for incidence, information about incidence is shared across time and the degradation in overall inference performance (as assessed by CRPS) is generally minor (Fig. S4).

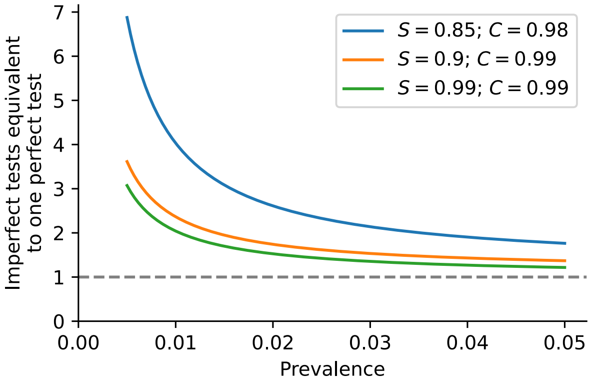


Figure S6: **The number of imperfect tests achieving the same Fisher information for actual prevalence as a single perfect test.** For three selected sensitivity and specificity values, we plot the equivalent number of imperfect tests as computed in eq. (6) as a function of actual prevalence.
